# Health Care Utilization of US Medicare Beneficiaries Living with Parkinson’s Disease

**DOI:** 10.1101/2022.06.03.22275470

**Authors:** Caroline Pearson, Alex Hartzman, Dianne Munevar, Megan Feeney, Rachel Dolhun, Veronica Todaro, Sheera Rosenfeld, Allison Willis, James Beck

**Affiliations:** NORC at the University of Chicago; Parkinson’s Foundation; Michael J. Fox Foundation; University of Pennsylvania Perelman School of Medicine

## Abstract

The number of people living with Parkinson’s disease (PD) is expected to rise in the coming years. This study analyzed health care utilization patterns of Medicare beneficiaries with a PD diagnosis (ICD-10 code G20) who were enrolled in 2019. Utilization analysis included PD-related specialists and primary care physicians, therapy services, and mental health services. We found 685,116 (1.2%) Medicare beneficiaries had PD (56.3% male, 77.9% over age 70, 85.3% White, and 16.0% rural residents). Few Medicare beneficiaries with PD sought care from a movement disorder specialist (MDS) (9.1%); another 50.9% visited a general neurologist. Results reveal low utilization rates for therapy and even lower rates for mental health services. Overall healthcare utilization varied significantly by demographic group with women, people of color, and rural residents being less likely to access specialist care. Our findings emphasize the need for further research on population-specific barriers to accessing PD-related health care.

## Introduction

Within the United States, an estimated 89% of those diagnosed with PD are eligible for Medicare either because of their age (65 years of age and older) or prolonged disability status. [1] However, few studies have examined the health care utilization patterns of people living with PD on Medicare and how demographic differences, especially for groups that have been historically underrepresented in research, impact utilization of health care services. [2, 3]

Inconsistent symptom presentation and disease progression, as well as lack of biomarker or objective clinical diagnostic test to diagnose disease creates a challenge for diagnosing and treating PD, especially for physicians with less expertise in movement disorders. [4] As such, diagnosis and care is typically managed by general neurologists and/or movement disorder specialists in an outpatient setting. [5, 6] Previous research finds that movement disorder specialists more accurately diagnose PD, especially in earlier stages and atypical presentations, than neurologists without subspecialty training; benefits of early detection can include reduced risk of disease progression and improved quality of life. [7] Additionally, PD specialist involvement in the management of PD patient care has been found to improve the patient experience in all care settings and stages of care. [6] Given the importance of both motor and non-motor symptoms of PD, treatment for PD should include pharmaceutical interventions, along with rehabilitative and maintenance therapy, mental health services. [8]

As part of a broader portfolio of research on how people living with PD access health care and information, this analysis uses Medicare program data to explore the demographic characteristics, including gender, age, race and ethnicity, and rural residency (“urbanicity”), and PD-related health care utilization of Medicare beneficiaries living with PD using comprehensive data on Medicare beneficiaries and services made available by the Centers for Medicare & Medicaid Services. The implications of this study are particularly important when considered along with recent growth rates of incident Parkinson’s disease in the US population, which increased more than 50 percent over the past decade (2012-2020), as well as the increase in the American population and associated projected growth of the Medicare population, which is expected to grow by 20.3% between 2021 and 2029. [9, 10, 11]

## Results

### Prevalence and Demographics of Parkinson’s Disease in the Medicare Population

In 2019, there were 64,430,729 beneficiaries within the US Medicare system (Table 1). We restricted our analysis to those beneficiaries who had at least one claim with an ICD-10 diagnostic code of G20 indicating Parkinson’s disease (808,107 beneficiaries, comparable to prior Medicare-specific projections and age proxied U.S. and international projections. [1, 12, 13, 14]). Subsequently, we limited analyses further to beneficiaries who were continuously enrolled in full coverage Medicare Fee-for-Service (FFS) (Parts A and B) or a Medicare Advantage (Part C) plan during the 2019 calendar year and had observable physician information included in their claims or encounter records. This identified 685,116 or 1.2% of the corresponding total Medicare population with at least one ICD-10 code diagnosis of “Parkinson’s disease”.

**Table 1.** Exclusion criteria for study population, “2019 Medicare Parkinson’s disease population,” 2019

|  |  |  |
| --- | --- | --- |
|  | **Excluded**<br>*n* | **Remaining**<br>*n* |
| 1. Beneficiaries enrolled in 2019 | - | 64,430,729 |
| 2. Beneficiaries with at least 1 G20 (“Parkinson’s Disease”) diagnosis in 2019 | 63,622,622 | 808,107 |
| 3. Beneficiary deaths in 2019 | 80,423 | 727,684 |
| 4. Beneficiaries enrolled for only part of 2019 | 8,698 | 718,986 |
| 5. Beneficiaries with partial FFS/MA coverage in 2019 | 19,870 | 699,116 |
| 6. Beneficiaries missing provider information in all Medicare claims in 2019 | 14,000 | 685,116 |
| **2019 Medicare beneficiaries with Parkinson’s Disease (study population)** |  | **685,116** |

People living with PD enrolled in Medicare tended to be older relative to the Medicare population as a whole (77.9% versus 57.1% over age 70, respectively). Medicare beneficiaries with PD were also more likely to be male (56.3%) than the broader Medicare population, which is 45.6% male and 54.4% female. The PD population was only slightly less likely to reside in rural areas (16.0%) compared to other Medicare beneficiaries (17.5%). 39.5% of beneficiaries living with PD were enrolled in Medicare Advantage plans compared to 37.3% of Medicare beneficiaries overall.

People living with PD were 2.3% Asian, 5.9% Black, 2.6% Hispanic, 0.3% North American Native, and 85.3% White (Table 2). People of color tend to be underrepresented among beneficiaries living with PD relative to the overall Medicare population; whether this is due to underdiagnosis or truly lower prevalence of PD among these populations is not known.

**Table 2.**
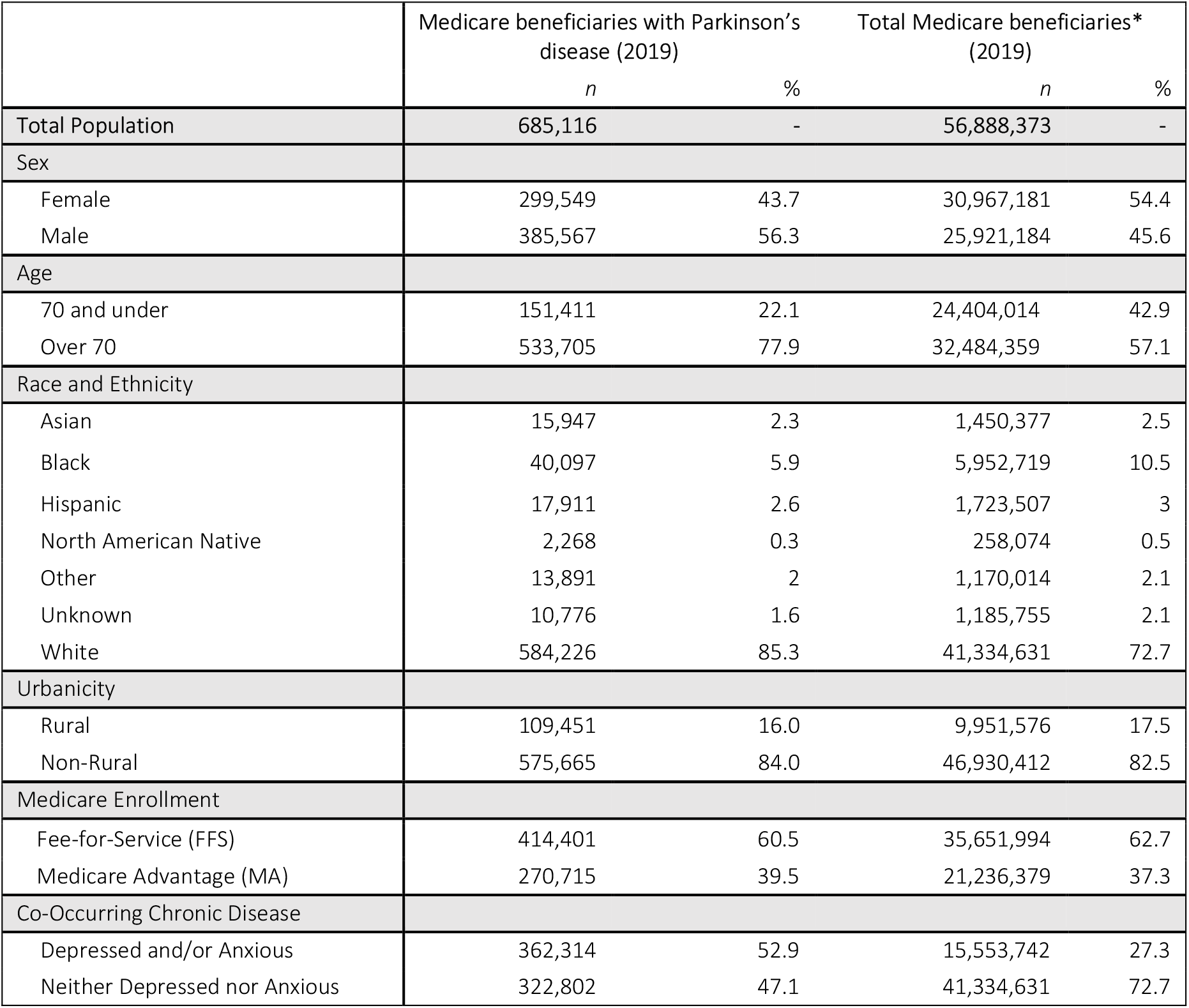
Prevalence of Parkinson’s disease diagnosis by demographic characteristics in Medicare, 2019

|  |  |  |  |  |
| --- | --- | --- | --- | --- |
|  | **Medicare beneficiaries with Parkinson’s disease (2019)** |  | **Total Medicare beneficiaries\* (2019)** |  |
|  | *n* | % | *n* | % |
| **Total Population** | **685,116** | **-** | **56,888,373** | **-** |
| **Sex** |  |  |  |  |
| Female | 299,549 | 43.7 | 30,967,181 | 54.4 |
| Male | 385,567 | 56.3 | 25,921,184 | 45.6 |
| **Age** |  |  |  |  |
| 70 and under | 151,411 | 22.1 | 24,404,014 | 42.9 |
| Over 70 | 533,705 | 77.9 | 32,484,359 | 57.1 |
| **Race and Ethnicity** |  |  |  |  |
| Asian | 15,947 | 2.3 | 1,450,377 | 2.5 |
| Black | 40,097 | 5.9 | 5,952,719 | 10.5 |
| Hispanic | 17,911 | 2.6 | 1,723,507 | 3 |
| North American Native | 2,268 | 0.3 | 258,074 | 0.5 |
| Other | 13,891 | 2 | 1,170,014 | 2.1 |
| Unknown | 10,776 | 1.6 | 1,185,755 | 2.1 |
| White | 584,226 | 85.3 | 41,334,631 | 72.7 |
| **Urbanicity** |  |  |  |  |
| Rural | 109,451 | 16.0 | 9,951,576 | 17.5 |
| Non-Rural | 575,665 | 84.0 | 46,930,412 | 82.5 |
| **Medicare Enrollment** |  |  |  |  |
| Fee-for-Service (FFS) | 414,401 | 60.5 | 35,651,994 | 62.7 |
| Medicare Advantage (MA) | 270,715 | 39.5 | 21,236,379 | 37.3 |
| **Co-Occurring Chronic Disease** |  |  |  |  |
| Depressed and/or Anxious | 362,314 | 52.9 | 15,553,742 | 27.3 |
| Neither Depressed nor Anxious | 322,802 | 47.1 | 41,334,631 | 72.7 |

### Health Care Utilization – Physician Services

The majority (60.0%) of Medicare beneficiaries with PD had at least one visit with a neurology specialist (i.e., general neurologist or a movement disorder specialist) in 2019. 40% instead sought care from a primary care physician or did not see a physician for their PD at all during the year (Table 3). Specifically, only 9.1% of individuals with PD visited an MDS at least once in 2019 (Table 3). 50.9% of the population did not have an MDS visit but had at least one visit with a general neurologist (MDS=0, GN ≥1) (Table 3).

**Table 3.**
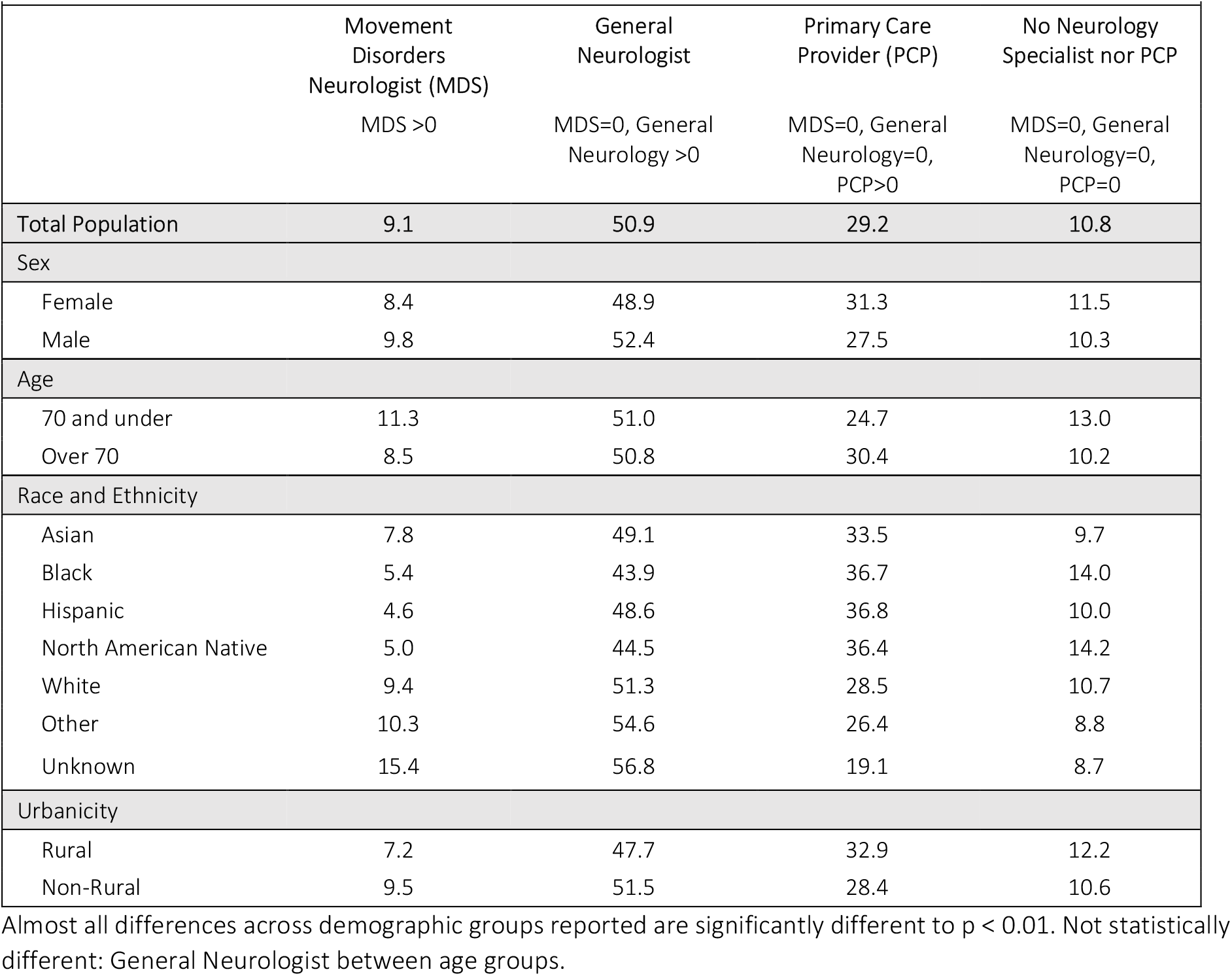
Utilization of physician services by Medicare beneficiaries with PD, by demographic characteristics, 2019 (percent)

|  |  |  |  |  |
| --- | --- | --- | --- | --- |
|  | **Movement Disorders Neurologist (MDS)**<br>MDS >0 | **General Neurologist**<br>MDS=0, General Neurology >0 | **Primary Care Provider (PCP)**<br>MDS=0, General Neurology=0, PCP>0 | **No Neurology Specialist nor PCP**<br>MDS=0, General Neurology=0, PCP=0 |
| **Total Population** | **9.1** | **50.9** | **29.2** | **10.8** |
| **Sex** |  |  |  |  |
| Female | 8.4 | 48.9 | 31.3 | 11.5 |
| Male | 9.8 | 52.4 | 27.5 | 10.3 |
| **Age** |  |  |  |  |
| 70 and under | 11.3 | 51.0 | 24.7 | 13.0 |
| Over 70 | 8.5 | 50.8 | 30.4 | 10.2 |
| **Race and Ethnicity** |  |  |  |  |
| Asian | 7.8 | 49.1 | 33.5 | 9.7 |
| Black | 5.4 | 43.9 | 36.7 | 14.0 |
| Hispanic | 4.6 | 48.6 | 36.8 | 10.0 |
| North American Native | 5.0 | 44.5 | 36.4 | 14.2 |
| White | 9.4 | 51.3 | 28.5 | 10.7 |
| Other | 10.3 | 54.6 | 26.4 | 8.8 |
| Unknown | 15.4 | 56.8 | 19.1 | 8.7 |
| **Urbanicity** |  |  |  |  |
| Rural | 7.2 | 47.7 | 32.9 | 12.2 |
| Non-Rural | 9.5 | 51.5 | 28.4 | 10.6 |

Almost all differences across demographic groups reported are significantly different to $p < 0.01$ . Not statistically different: General Neurologist between age groups.

There were significant demographic differences in physician utilization. As shown in Table 3, female beneficiaries living with PD seek care from MDS and general neurologists at lower rates than male beneficiaries. 8.4% of female beneficiaries living with PD visited an MDS at least once in 2019 compared with 9.8% of male beneficiaries. 48.9% of female beneficiaries that did not have an MDS visit in 2019 had at least one general neurology visit compared to 52.4% of male beneficiaries (Table 3). Medicare beneficiaries under the age of 70 were more likely to have had at least one MDS visit in 2019 (11.3%) than older beneficiaries (8.5%). Reported differences are significant to p < 0.01; please see tables for further detail.

Asian, Black, Hispanic, and North American Native beneficiaries with PD utilized specialty care at lower rates than White beneficiaries. 7.8% of Asian beneficiaries, 5.4% of Black beneficiaries, 4.6% of Hispanic beneficiaries, and 5.0% of North American Native beneficiaries had at least one MDS visit in 2019 compared to 9.4% of White beneficiaries. 51.3% of White beneficiaries did not use MDS care but had at least one general neurology visit compared with 49.1% of Asian beneficiaries 43.9% of Black beneficiaries, 48.6% of Hispanic beneficiaries, and 44.5% North American Native beneficiaries (Table 3).

Individuals residing in rural areas were less likely to receive care from an MDS than their urban counterparts. Only 7.2% of rural residents saw an MDS at least once in 2019, compared to 9.5% of urban residents. Rural residents were also less likely than non-rural residents to visit a general neurologist if they did not visit an MDS (47.7% of rural residents vs. 51.5% of non-rural residents) (Table 3).

### Health Care Utilization – Therapy and Mental Health Services

Since treating the motor symptoms of PD is crucial to manage the disease, we examined the number of PD Medicare beneficiaries that used physical, occupational, and speech-language therapy providers (Table 4) where PD was listed on the claim. In 2019, 20.3% of the population of Medicare beneficiaries living with PD used physical therapy, 9.5% used occupational therapy, and 7.5% used speech-language therapy. Additionally, despite 32.8% of Medicare beneficiaries with PD having a diagnosis of depression and/or anxiety, only 2.1% of these individuals had at least one clinical psychology visit and 4.4% had at least one psychiatry visit (see Supplemental Table 2).

**Table 4.** Percent utilization of therapy services by Medicare beneficiaries with PD, by demographic characteristics, 2019

|  |  |  |  |
| --- | --- | --- | --- |
|  | Physical Therapy | Occupational Therapy | Speech-Language Therapy |
| **Total Population** | **20.3** | **9.5** | **7.5** |
| Sex |  |  |  |
| Female | 19.8 | 10.2 | 6.9 |
| Male | 20.6 | 8.9 | 7.9 |
| Age |  |  |  |
| 70 and under | 17.3 | 7.3 | 6.2 |
| Over 70 | 21.1 | 10.1 | 7.8 |
| Race and Ethnicity |  |  |  |
| Asian | 14.9 | 5.8 | 5.1 |
| Black | 16.9 | 9.7 | 6.7 |
| Hispanic | 12.3 | 5.4 | 4.2 |
| North American Native | 18.9 | 7.9 | 6.7 |
| **White** | **20.9** | **9.8** | **7.7** |
| Other | 19.4 | 6.9 | 6.3 |
| Unknown | 22.0 | 8.1 | 7.5 |
| Urbanicity |  |  |  |
| Rural | 19.3 | 9.6 | 7.4 |
| Non-Rural | 20.5 | 9.5 | 7.5 |

Almost all differences across demographic groups reported are significantly different to $p < 0.01$ . Not statistically different: Occupational therapy between Black and White, Physical therapy and Speech-Language therapy between North American Native and White, Speech-Language therapy between ‘unknown’ race/ethnicity and White, and Occupational therapy and Speech-Language therapy between rural and non-rural urbanicity.

There were significant demographic differences in utilization of physical, occupational, and speech-language therapy among Medicare beneficiaries living with PD, shown in Table 4. Male beneficiaries with PD were more likely to use physical therapy (20.6%) or speech-language therapy (7.9%) than female beneficiaries (19.8% and 6.9%, respectively) (Table 4). Individuals over age 70 are more likely to use therapy services than those under the age of 70 (Table 4). Asian, Hispanic, and North American Native individuals used all therapy services at rates below average. 9.7% of Black beneficiaries used occupational therapy, a higher rate than the 9.5% average. Rural and non-rural Medicare beneficiaries living with PD accessed therapy services at similar rates (Table 4).

Medicare beneficiaries with PD who received specialist care during the year were generally more likely to use therapy and mental health services, with those seeing an MDS being the most likely to access these services (Table 5 and Supplemental Table 2). Medicare beneficiaries who had at least one MDS visit in 2019 were most likely to use therapy and mental health service providers in the same year – 13.1% used occupational therapy, 33.2% used physical therapy, 13.1% used speech-language therapy, 3.8% used clinical psychology, and 3.7% used psychiatry (Table 5 and Supplemental Table 2). Individuals who did not see an MDS but used general neurology services at least once per year were more likely to used physical and speech-language therapy and clinical psychology services than those who did not seek care from neurology specialists (i.e., either an MDS or general neurologist) (Table 5 and Supplemental Table 2).

**Table 5.** Percent utilization of therapy services as a function of physician utilization, 2019

|  |  |  |  |
| --- | --- | --- | --- |
|  | Occupational Therapy | Physical Therapy | Speech-Language Therapy |
|  | % | % | % |
| **MDS** |  |  |  |
| *MDS >0* | 13.1 | 33.2 | 13.1 |
| **General Neurology** |  |  |  |
| *MDS=0, General Neurology >0* | 9.2 | 23.2 | 7.7 |
| **PCP** |  |  |  |
| *MDS=0, General Neurology=0, PCP>0* | 11.4 | 16.3 | 7.4 |
| **None** |  |  |  |
| *MDS=0, General Neurology=0, PCP=0* | 2.6 | 6.4 | 1.7 |

All differences across primary provider utilization are significantly different to $p < 0.01$ .

## Discussion

### Prevalence and Demographics of Parkinson’s Disease in the Medicare Population

This study uses large-scale administrative data on Parkinson’s disease prevalence from the most inclusive, population-based U.S. health care database to identify the size, demographic characteristics, and health care utilization patterns of the population of Medicare beneficiaries living with PD. An important strength of this study is the substantial size of the Medicare data set, including the totality of the Fee-for-Service and Medicare Advantage populations included in a single study. The use of this data as the study set allows for a robust analysis of the demographic characterization of the Medicare beneficiary population living with PD, and more reliable estimates of health care utilization than previous studies that rely on data collected from hospitals and physician groups or limited solely to Medicare FFS. Additionally, past research has largely focused on the prevalence of PD in White people, males, and those living in non-rural areas. [15, 2, 3] In comparison, this study analyzed a population of over 685,000 Medicare beneficiaries, reducing bias related to underrepresentation that is more prevalent in survey research and in location-specific utilization research. [3]

We also find that male beneficiaries were 1.22 times more likely to have PD than female beneficiaries. Several studies find that PD is more common in men than women, by male-to-female ratios ranging between 1.1 and 2.7 in populations 50 years of age and older. [16] A previous study of the Medicare population found PD was 1.39 times more prevalent in male than female beneficiaries. [17]

Prevalence estimates in our study for the total Medicare population is likely underestimated given that estimates are based on a one-year period (2019) in which individuals must have been continuously enrolled, had full Medicare coverage, and sought care from a provider that recorded their PD diagnosis on at least one billed outpatient or physician office service claim. Therefore, individuals who did not seek care from a Medicare provider are not captured in these estimates. Additionally, prevalence was underestimated in cases where diagnoses were incompletely or inaccurately captured in claims, or among individuals who had interruptions or changes to their Medicare enrollment or coverage.

### Health Care Utilization – Physician Services

Our study finds that access to and utilization of specialist physician services among individuals living in rural areas is significantly lower than utilization for those living in non-rural areas. 7.2% of rural residing Medicare beneficiaries utilized MDS care, compared to 9.5% of non-rural residents and 9.1% of all beneficiaries with PD. Utilization of general neurology care among those who did not utilize MDS care is also lower for rural residents (47.7%) than non-rural residents (51.5%). This may be explained by access barriers related to distance from PD specialists which studies suggest reduces utilization of PD specialty care, particularly for lower-income populations and those living in rural areas. [3, 6] And distance to neurology specialists, particularly MDS, is not the only limiting factor—there are about 660 MDS in the US, which translates to an average of one MDS per 1,038 Medicare beneficiaries living with PD. However, for rural residents, distance and access to MDS is even more challenging since only 6 out of 660 MDS practice in rural areas. Despite disparities in accessing specialist care, individuals with PD living in rural areas received therapy services at similar rates to people in non-rural areas, which may suggest specialist shortages in rural areas, but adequate referrals to therapy providers from primary care providers.

Individuals under the age of 70 were more likely to utilize specialty care and less likely to use therapy services – this may be due to the prevalence of newer diagnoses in the population under 70 compared to those over 70, which is confirmed in various studies on population-level PD onset. [12, 18, 14] Individuals earlier in their disease progression are more likely to be in the diagnostic phase of their disease which requires multiple specialist evaluations and may include testing various supplemental care offerings to find the combination of care that best manages their symptoms. [5, 7]

We find that 32.8% of Medicare beneficiaries with PD have a diagnosis of depression and/or anxiety, but only 2.1% of these individuals had at least one clinical psychology visit and 4.4% had at least one psychiatry visit. This may be because there is a gap in mental health coverage in current Medicare policies which creates cost and network barriers for beneficiaries attempting to access needed mental health services. [19] Due to the lack of Medicare insurance coverage of mental health services, beneficiaries may seek and pay for mental or behavioral health services out-of-pocket, which is thereby not captured in the administrative claims data used for this analysis.

### Health Care Utilization – Therapy and Mental Health Services

Although the use of therapy services such as physical, occupational therapy, and speech-language therapy and mental health services are considered key interventions for the management of PD, utilization of these services remains low among Medicare beneficiaries living with PD. Our study finds that utilization of specialist physician care is a predictor of utilization of therapy and mental health services. There were significant differences in utilization of therapy and mental health providers depending on the type of physician from which an individual sought care. Medicare beneficiaries who visited an MDS in 2019 were most likely to also have at least one physical therapy visit (33.2%), occupational therapy visit (13.1%), and/or speech-language therapy visit (13.1%). Utilization of mental health services for Medicare beneficiaries who get PD care from an MDS shows a similar pattern—3.8% had a clinical psychology visit, 3.7% had at least one psychiatry visit. It is likely that this is because MDS are specifically trained to coordinate care for their patients with PD, and studies have found that integration of an MDS in all stages and settings of care is associated with an improvement in patient experience and quality of life. [4, 5, 6, 7]

There are potential causes for both under- and over-estimation of the PD services utilization among the population in this study. First, this study likely underestimates utilization of health care services in the population of Medicare beneficiaries living with PD, due to missing provider data, inaccurate and/or incomplete coding, and interruptions or changes to Medicare enrollment and/or coverage. However, a source of potential overestimation is that we assume that individuals living with PD are seeking care from MDS and general neurologists primarily for the management of their PD as a PD diagnosis (ICD-10-CM code G20) must be included on the claim for us to measure it; that said, this study does not explicitly account for co-existing conditions or services rendered during physician visits. This overestimation effect is likely greater when considering therapy given that physical and occupational therapy, and speech-language therapy are commonly used to manage other conditions that are common among older adults (noting all utilization measured in this study had to include a PD diagnosis on the claim).

## Conclusion

This analysis reveals significant gaps in how many Medicare beneficiaries with PD access health care, compared to recommended best practice. It further reveals persistent health disparities for women, people of color, and rural residents—each of whom may face challenges with PD diagnosis and access to treatment. Finally, the analysis demonstrates differences in referral patterns among physician types, with patients seen an MDS being more likely to use therapy and mental health services to treat the common symptoms of PD, indicating these clinicians may serve patients with more advanced PD. This suggests opportunities to improve health equity and quality of care by expanding PD-specific training for general neurologists and pursuing strategies to improve access to care across demographic groups and geographies. PCPs might be supported by encouraging referrals to MDS and general neurologists at the time of a suspected PD diagnosis.

## Materials and Methods

This retrospective observational study assesses the prevalence of Parkinson’s disease in the 2019 Medicare population, and healthcare utilization of Medicare beneficiaries living with Parkinson’s disease.

### Data

This study was conducted using 2019 Medicare enrollment data, Fee-for-Service claims (Parts A and B), and Medicare Advantage (Part C) encounter data obtained from the Centers for Medicare & Medicaid Services (CMS). Demographic and location data was retrieved from Medicare Beneficiary Summary Files. Diagnostic, provider specialty, and utilization information was derived from administrative claims and encounter data. Some Part C encounter data lacked information on provider identity or specialty; where possible this was imputed from data available in Part A and B claims. Beneficiaries enrolled in Part C whose encounter data were entirely lacking provider information were excluded from demographics and utilization measures.

### Prevalence

Prevalence of Parkinson’s disease in the Medicare population during the study year, 2019, is defined as the number of Medicare beneficiaries continuously enrolled in the same calendar year with one or more Medicare claims with a primary or secondary diagnosis of PD. Parkinson’s disease was identified using International Classifications of Diseases, 10^th^ Revision, Clinical Modification (ICD-10-CM) code G20. Medicare beneficiaries with partial Medicare FFS (Parts A and B) or Medicare Advantage (Part C) coverage, and beneficiaries with missing provider information in all medical claims were not included in the study. ICD-10 coding for Parkinson’s disease is more specific than ICD-9 codes used in prior studies and does not include secondary Parkinsonism diagnoses. [20] Of those identified with PD, 83.8% of beneficiaries had more than one medical service claim with a PD diagnosis in 2019 and 72.1% had more than two claims; demographics differ based on the number of applicable diagnostic codes with Black, Hispanic, and rural residents being more likely to have exactly one G20 code in the year. (Supplemental Table 1). We opted for the broadest possible inclusion criteria of at least one G20 code to include racial and ethnic groups and rural residents that may experience greater barriers to accessing PD care, but this definition may include some individuals who are undergoing testing but do not have an actual PD diagnosis in our results.

Of note, our identified Medicare prevalence of 685,116 (after exclusions), about 1.2% of the total population, is consistent to the approximately 450,000 observed in the 2000-2005 Medicare FFS dataset by Willis et al (which was 1.6% of the FFS population at the time). [21] Also like Willis et al. [21], we observe that Asian, Black, and Hispanic people tend to be less well-represented among PD patients than the overall Medicare population. It is not known if this is indicative of disparities in screening and diagnosis of PD among these racial and ethnic groups, or if it points to a potential genetic or environmental component of PD that more often affects White people. Previous research confirms that PD is most commonly found in White people, but also finds that racial and ethnic differences in incident PD cannot be explained by differences in age, sex, income, insurance, or healthcare utilization. Therefore, research suggests that the prevalence of PD may be explained by biological differences, environmental factors, or other social determinants. [21, 17] Research also suggests that Black and Hispanic people are less likely to use health care services or self-report symptoms than White people, which may also contribute to underdiagnosis in these populations. [22, 23, 2]

### Healthcare Services Utilization

Utilization was calculated for physician services (neurology – movement disorder specialist, general neurology, and primary care), therapy services (physical, occupational, and speech-language therapy), and mental health services (clinical psychology and psychiatry). Healthcare utilization was summarized as a function of age, sex, race and ethnicity, and rurality derived from data found in the Master Beneficiary Summary File. Chronic condition flags for anxiety and depression indicators were applied for FFS enrollees; these flags were reconstructed using diagnostic information on encounter data for MA enrollees.

Additionally, therapy utilization and mental health service utilization was analyzed as a function of physician service utilization.

### Statistical Analyses

Demographic characteristics and prevalence of Parkinson’s disease in the Medicare population were summarized with descriptive statistics. Inferential statistics were used to describe and quantify cohort differences in healthcare services utilization. Outcomes were compared statistically between cohorts using chi-squared tests. Differences between cohorts were considered statistically significant at p<0.01. Excel software was used for statistical analysis.

## Data Availability

Data supporting this study are not publicly available. Due to CMS policy, the authors are unable to provide the data used in this study. Data can be requested through ResDAC’s CMS Data Request Center.

https://resdac.org/

## Acknowledgements

We would like to thank Parkinson’s Foundation Christiana Evers, Vice President and Chief Community Engagement Officer and Nicole Yarab, Vice President, Clinical Affairs and Information & Resources, Parkinson’s Foundation Research Advocates, Lisa Seghetti and A.C. Woolnough, as well as Web Ross, MD, Pacific Health Research and Education Institute, for their helpful review of the manuscript. We are also grateful for the research support provided by Amrithaa Gunabalan and Morgan Clausen, statistical programming from Kevin Dietz, and technical assistance from Ryan Murphy. Data used in the preparation of this article were obtained from The Centers for Medicare & Medicaid Services (CMS) through a Research Data Use Agreement (DUA) with NORC at the University of Chicago. This research was supported by the Parkinson’s Foundation and the Michael J. Fox Foundation.

