## Supplemental Tables for "Health Care Utilization of US Medicare Beneficiaries Living with Parkinson’s Disease"

**Supplemental Table 1.** Demographics of Medicare Enrollees by Number of Claims with a PD (G20) ICD-10 Diagnostic Code, 2019

| | At Least 1 G20 Dx<br>% | At Least 2 G20 Dx<br>% | Exactly 1 G20 Dx<br>% |
|---|---|---|---|
| Sex | | | |
| Female | 43.8 | 43.4 | 47.1 |
| Male | 56.2 | 56.6 | 52.9 |
| Age | | | |
| 70 and under | 22.1 | 21.6 | 26.1 |
| Over 70 | 77.9 | 78.4 | 73.9 |
| Race and Ethnicity | | | |
| Asian | 2.3 | 2.3 | 2.4 |
| Black | 5.9 | 5.7 | 7.7 |
| Hispanic | 2.7 | 2.6 | 2.9 |
| North American Native | 0.3 | 0.3 | 0.4 |
| Other | 2.0 | 2.0 | 1.8 |
| Unknown | 1.6 | 1.6 | 1.4 |
| White | 85.2 | 85.4 | 83.4 |
| Urbanicity | | | |
| Rural | 15.9 | 15.7 | 17.7 |
| Non-Rural | 84.1 | 82.2 | 82.3 |

**Supplemental Table 2.** Utilization of Mental Health Services as a Function of Physician Utilization, 2019

| | Clinical Psychology<br>% | Psychiatry<br>% |
|---|---|---|
| MDS<br>*MDS >0* | 3.8 | 3.7 |
| General Neurology<br>*MDS=0, General Neurology >0* | 1.5 | 2.3 |
| PCP<br>*MDS=0, General Neurology=0, PCP>0* | 0.6 | 2.2 |
| None<br>*MDS=0, General Neurology=0, PCP=0* | 0.7 | 3.14 |
